# Tenofovir Disoproxil Fumarate and severity of COVID-19 in people with HIV infection

**DOI:** 10.1101/2021.11.11.21266189

**Authors:** J Del Amo, R Polo, S Moreno, E Martínez, A Cabello, JA Iribarren, A Curran, J Macías, M Montero, C Dueñas, AI Mariño, S Pérez de la Cámara, A Díaz, JR Arribas, I Jarrín, MA Hernán, on behalf of the CoVIHd Collaboration in Spain

## Abstract

**Background:** Effective, safe, and affordable antivirals are needed for COVID-19. Tenofovir has not been studied in randomized trials despite evidence consistent with its effectiveness against COVID-19.

**Methods:** We studied HIV-positive individuals on antiretroviral therapy (ART) in 2020 at 69 HIV clinics in Spain. We collected data on sociodemographics, ART, CD4-cell count, HIV-RNA viral load, comorbidities and the following outcomes: laboratory-confirmed SARS-CoV-2 infection, COVID-19 hospitalization, intensive care unit (ICU) admission and death. We compared the 48-week risks for individuals receiving tenofovir disoproxil fumarate (TDF)/emtricitabine (FTC), tenofovir alafenamide (TAF)/ FTC, abacavir (ABC)/lamivudine (3TC), and other regimes. All estimates were adjusted for clinical and sociodemographic characteristics via inverse probability weighting.

**Results:** Of 51,558 eligible individuals, 39.6% were on TAF/FTC, 11.9% on TDF/FTC, 26.6% on ABC/3TC, 21.8% on other regimes. There were 2,402 documented SARS-CoV-2 infections (425 hospitalizations, 45 ICU admissions, 37 deaths). Compared with TAF/FTC, the estimated risk ratios (RR) (95% CI) of hospitalization were 0.66 (0.43, 0.91) for TDF/FTC and 1.29 (1.02, 1.58) for ABC/3TC, the RRs of ICU admission were 0.28 (0.11, 0.90) for TDF/FTC and 1.39 (0.70, 2.80) for ABC/3TC, and the RRs of death were 0.37 (0.23, 1.90) for TDF/FTC and 2.02 (0.88-6.12) for ABC/3TC. The corresponding RRs of hospitalization for TDF/FTC were 0.49 (0.24, 0.81) in individuals ≥50 years and 1.15 (0.59, 1.93) in younger individuals.

**Conclusion:** Our findings suggest that, compared with other antiretrovirals, TDF/FTC lowers COVID-19 severity among HIV-positive individuals with virological control. This protective effect may be restricted to individuals aged 50 years and older.

## Introduction

While vaccine rollout is ongoing, most of the world population remains vulnerable to COVID-19. Therefore, much research has focused on the repurposing of antivirals against SARS-CoV-2. Remdesivir, originally developed against the Ebola virus, was intensively studied from the start of the pandemic^1–3^. More recently molnupiravir, originally developed against the influenza virus, has been reported to reduce the risk of hospitalization and death in high-risk COVID-19 patients^4^.

The repurposing of tenofovir against SARS-CoV-2, however, has not been pursued further despite being a safe and affordable drug that may be effective for the prevention or treatment of COVID-19^5–9^. Three observational studies found lower risks of SARS-CoV-2 antibody prevalence^7^, symptomatic COVID-19^5^, COVID-19 hospitalization,^5^ or COVID-19 mortality^5,6^ among HIV-positive individuals receiving tenofovir disoproxil fumarate/emtricitabine (TDF/FTC) than among those receiving other antiretroviral regimes. Another observational study found a lower risk of severe COVID-19 in patients with chronic hepatitis B virus (HBV) infection treated with TDF/FTC than among those treated with entecavir^8^.

A key unanswered question is whether the association between TDF/FTC and lower risk of serious COVID-19 reflects a causally protective effect of TDF/FTC or simply confounding because TDF/FTC may be used in healthier individuals (e.g., patients with no renal disease) with a lower pre-existing risk of severe COVID-19^5,10^. Distinguishing between a true causal effect and confounding is difficult because no findings from randomized trials of TDF/FTC with clinical COVID-19 outcomes are yet available, and previous observational studies were either not based on a sufficiently large sample of individuals on antiretroviral therapy or did not adjust for a broad range of potentially important confounders.

Here we report the findings from a nationwide cohort study of TDF/FTC and COVID-19 outcomes among HIV-positive individuals on antiretroviral therapy with optimal virological control. We collected data on and adjusted for key potential confounders, including hypertension, diabetes, chronic renal disease, cardiovascular disease, and treatment with immunosuppressants or corticosteroids.

## Methods

### Study population

HIV-positive individuals in Spain receive care at specialized hospital outpatient clinics. The CoVIHd Collaboration (Covid-19 in HIV-positive individuals in Spain) includes HIV-positive individuals who were receiving antiretroviral therapy at the HIV clinics of 87 Spanish hospitals between January 1 and December 31, 2020. All clinics collected information on individuals with a history of SARS-CoV-2 infection, but this analysis is restricted to the 69 clinics that collected information on HIV-positive individuals with and without a history of SARS-CoV-2 infection. These 69 clinics serve approximately 44% of all persons on antiretroviral therapy with virological suppression in Spain^11^.

Hospitals transmitted de-identified data to the coordinating center at the Institute of Health Carlos III in Madrid via a secure web-based application specifically designed for this purpose. For each individual, data included sociodemographic characteristics, dates and composition of all antiretroviral therapy regimes received during the study period, latest CD4 cell count and HIV RNA measurements before a COVID-19 diagnosis, comorbidities (from medical records, see Appendix 2), and date of laboratory-confirmed documented diagnosis of SARS-CoV-2 infection defined as positive results from a polymerase chain reaction (PCR) test (or, in a minority of cases, a SARS-CoV-2 antigen test or antibody test), following the Ministry of Health protocols^12^. The ascertainment of hospitalizations due to COVID-19, intensive care unit (ICU) admissions due to COVID-19, and deaths from COVID-19 was complete, but no protocol was in place to systematically screen for asymptomatic infections and mild cases of COVID-19.

### Eligibility criteria and follow-up

We included HIV-positive individuals aged 18 years or older who on February 1, 2020 had not received a diagnosis of SARS-CoV-2 infection and were on antiretroviral therapy, and who had virologically suppression (HIV RNA less than 50 copies/ml) in 2020. Virological suppression is an indicator of adequate adherence to antiretroviral therapy. For each individual, follow-up started on February 1 and ended on December 31, 2020. The goal was to emulate a (hypothetical) target trial in which individuals are randomly assigned to a particular nucleos(t)ide reverse transcriptase inhibitor (NRTI) combination before the start of SARS-CoV-2 transmission in their communities.

### Antiretroviral therapy regimes

We classified antiretroviral therapy regimes according to their NRTI combination into 4 categories: tenofovir disoproxil fumarate (TDF)/emtricitabine (FTC), tenofovir alafenamide (TAF)/ FTC, abacavir (ABC)/lamivudine (3TC), or other drug regimens excluding TDF, TAF and ABC. Most of the other drugs category were dual therapies including only one NRTI (3TC) (Appendix Table 1). We also studied regimens with three drugs according to the third drug class used along with the NRTI combination: integrase inhibitor, protease inhibitor, or nonnucleoside reverse transcriptase inhibitor (NNRTI).

### Outcomes

The outcomes of interest were any documented laboratory-confirmed diagnosis of SARS-CoV-2 infection and progressively more severe subsets of COVID-19: hospitalization due to COVID-19, ICU admission due to COVID-19, and death due to COVID-19. In supplemental analyses, we also considered documented asymptomatic SARS-CoV-2 infections and mild COVID-19 that did not require hospitalization.

### Statistical Analysis

We calculated the 48-week risk (cumulative incidence) and 95% confidence interval (CI) for each outcome by NRTI combination. We estimated the risks using a pooled logistic model with indicators for NRTI combination (3 indicators, with TAF/FTC as the reference group), week of follow-up (linear and quadratic terms), and product terms between NRTI combination indicators and week of follow-up. To adjust for baseline prognostic factors, we used inverse probability (IP) weighting. To estimate the denominator of the weights we fit a multinomial logistic model for the four NRTI combinations with covariates: age (in years, linear and quadratic terms), sex (male, female), transmission category (heterosexual, homo/bisexual, injecting drug use, other), country of origin (Spain, other), CD4 (<350, 350-500, >500 cells/mm^3^), and indicators for hypertension, diabetes, chronic renal disease, cardiovascular disease, and treatment with immunosuppressants or corticosteroids. We compared the risks via risk differences and risk ratios and used a nonparametric percentile-based bootstrap with 500 samples to obtain 95% CIs.

To compare the risks by the non-NRTI drug in the antiretroviral regime, we fit a similar model with indicators for NNRTI, protease inhibitor, and integrase inhibitor. We conducted subgroup analyses by age group (<50, ≥50 years) and sex for documented SARS-CoV-2 infections, and COVID-19 hospitalization.

We also conducted several sensitivity analyses. To evaluate the potential impact of treatment changes regimes, we conducted an analysis in which individuals were censored if/when they switch from their baseline antiretroviral regime to another regime. To evaluate the potential impact of the choice of inverse probability weighting as the method to adjust for confounding, we repeated the analyses with adjustment via standardization and also estimated adjusted hazard ratios via a Cox regression model. To assess the impact of measured confounding due to comorbidities and other factors, we repeated the analysis with no adjustment at all. All analyses were conducted with Stata, version 15.0 (StataCorp).

This study was approved by the institutional review board at University Hospital Ramón y Cajal, Madrid, Spain.

## Results

Of 51,558 eligible individuals (Figure 1), 39.6% were receiving TAF/FTC, 11.9% TDF/FTC, 26.6% ABC/3TC, and 21.8% other regimes (see Appendix Table 1 for a description of regimes in each category). The baseline characteristics of individuals in each of the four groups defined by NRTI combination are shown in Table 1. Individuals receiving TDF/FTC and TAF/FTC had similar age, sex, and CD4 cell counts, and were slightly younger than those receiving ABC/3TC or other regimes. The proportion of injecting drug users was slightly lower in persons on TAF/FTC than in the other groups. Individuals in the TDF/FTC group had a lower prevalence of hypertension, diabetes, and chronic renal disease than individuals in the other groups.

**Table 1.** Baseline characteristics of 51,558 eligible individuals by NRTI combination in HIV-positive individuals, CoVIHd Collaboration, Spain, February-December 2020

|  | **TAF/FTC**<br>**N = 20,432**<br>**(39.6%)** | **TDF/FTC**<br>**N = 6,160**<br>**(11.9%)** | **ABC/3TC**<br>**N = 13,715**<br>**(26.6%)** | **Other regimes**<br>**N = 11,251**<br>**(21.8%)** |
| --- | --- | --- | --- | --- |
| **Sex [N (%)]** |  |  |  |  |
| **Men** | 16,527 (80.9) | 4,856 (78.8) | 10,797 (78.7) | 8,623 (76.6) |
| **Women** | 3,905 (19.1) | 1,304 (21.2) | 2,918 (21.3) | 2,628 (23.4) |
| **Age, years**<br>**[Median (IQR)]** | 49 (39 – 56) | 48 (39 – 55) | 51 (41 – 57) | 53 (45 – 58) |
| **Transmission category [N (%)]** |  |  |  |  |
| **Heterosexual contact** | 4,652 (22.8) | 1,463 (23.7) | 3,218 (23.5) | 2,726 (24.2) |
| **Homo/bisexual contact** | 8,939 (43.7) | 2,221 (36.1) | 4,859 (35.4) | 3,741 (33.2) |
| **Injecting drug use** | 3,501 (17.1) | 1,242 (20.2) | 2,665 (19.4) | 2,847 (25.3) |
| **Other** | 503 (2.5) | 147 (2.4) | 300 (2.2) | 328 (2.9) |
| **Unknown** | 2,837 (13.9) | 1,087 (17.6) | 2,673 (19.5) | 1,609 (14.3) |
| **Country of origin [N (%)]** |  |  |  |  |
| **Spain** | 12,632 (61.8) | 3,836 (62.3) | 8,553 (62.4) | 8,077 (71.8) |
| **Other** | 4,689 (22.9) | 1,367 (22.2) | 2,325 (16.9) | 1,568 (13.9) |
| **Unknown** | 3,111 (15.2) | 957 (15.5) | 2,837 (20.7) | 1,606 (14.3) |
| **CD4 cell count, cells/mm<sup>3</sup>** |  |  |  |  |
| **Median (IQR)** | 704 (509-933) | 700 (511-929) | 746 (536-994) | 718 (520-948) |
| **<350** | 2,059 (10.1) | 619 (10.0) | 1,179 (8.6) | 1,014 (9.0) |
| **350-500** | 2,775 (13.6) | 834 (13.5) | 1,735 (12.6) | 1,504 (13.4) |
| **>500** | 15,414 (75.4) | 4,671 (75.8) | 10,718 (78.1) | 8,620 (76.6) |
| **Unknown** | 184 (0.9) | 36 (0.6) | 83 (0.6) | 113 (1.0) |
| **Hypertension [N (%)]** |  |  |  |  |
| **No** | 16,804 (82.2) | 5,388 (87.5) | 10,897 (79.4) | 8,469 (75.3) |
| **Yes** | 3,091 (15.1) | 695 (11.3) | 2,589 (18.9) | 2,564 (22.8) |
| **Unknown** | 537 (2.6) | 77 (1.2) | 229 (1.7) | 218 (1.9) |
| **Diabetes [N (%)]** |  |  |  |  |
| **No** | 18,490 (90.5) | 5,707 (92.6) | 12,217 (89.1) | 9,736 (86.5) |
| **Yes** | 1,486 (7.3) | 380 (6.2) | 1,302 (9.5) | 1,305 (11.6) |
| **Unknown** | 456 (2.2) | 73 (1.2) | 196 (1.4) | 210 (1.9) |
| **Chronic renal disease [N (%)]** |  |  |  |  |
| **No** | 19,375 (94.8) | 5,952 (96.6) | 12,570 (91.6) | 10,028 (89.1) |
| **Yes** | 1,057 (5.2) | 208 (3.4) | 1,145 (8.3) | 1,223 (10.9) |
| **Cardiovascular disease [N (%)]** |  |  |  |  |
| **No** | 16,628 (81.4) | 5,511 (89.5) | 11,759 (85.7) | 9,568 (85.0) |
| **Yes** | 1,051 (5.1) | 302 (4.9) | 773 (5.6) | 887 (7.9) |
| **Unknown** | 2,753 (13.5) | 347 (5.6) | 1,183 (8.6) | 796 (7.1) |
| **Treatment with**<br>**immunosuppressants or**<br>**corticosteroids [N (%)]** |  |  |  |  |
| **No** | 13,631 (66.7) | 4,313 (70.0) | 8,768 (63.9) | 7,805 (69.4) |
| **Yes** | 174 (0.8) | 78 (1.3) | 163 (1.2) | 142 (1.3) |
| **Unknown** | 6,627 (32.4) | 1,769 (28.7) | 4,784 (34.9) | 3,304 (29.4) |

**Figure 1.**
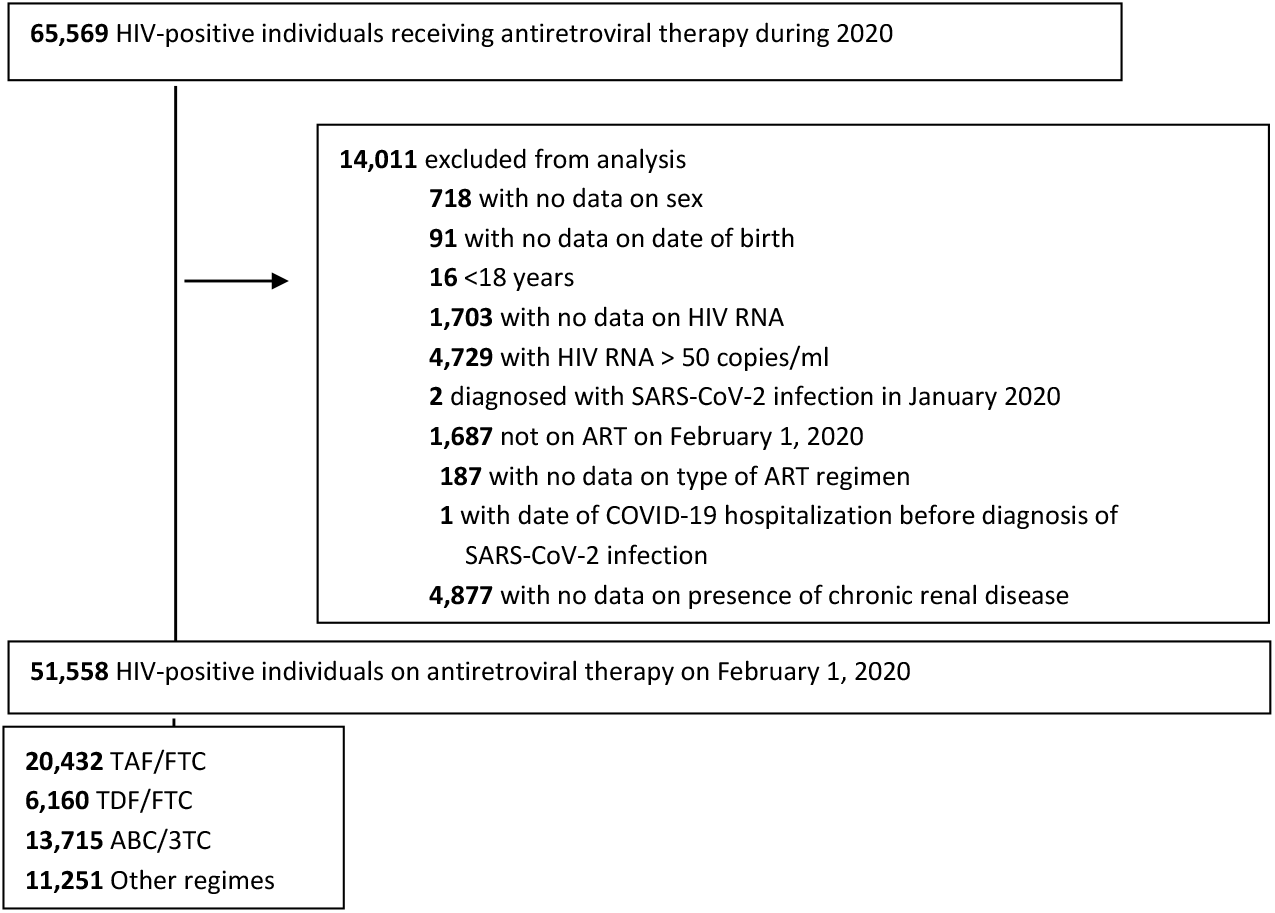
Flowchart of study population among HIV-positive individuals, CoVIHd Collaboration, Spain, February-December 2020

During the 48-week follow-up, there were 2,402 documented SARS-CoV-2 infections (425 hospitalizations, 45 ICU admissions, and 37 deaths). Of the 1,955 SARS-CoV-2 infections with available information on disease severity, 539 were asymptomatic, 1037 had mild COVID-19, 298 moderate COVID-19 and 81 severe COVID-19. Figure 2 shows the estimated cumulative risks of documented SARS-CoV-2 infection, COVID-19 hospitalization, COVID-19 ICU admission, and COVID-19 death by NRTI combination. Appendix Figure 1 shows the risks of asymptomatic COVID-19 and mild COVID-19.

**Figure 2.**
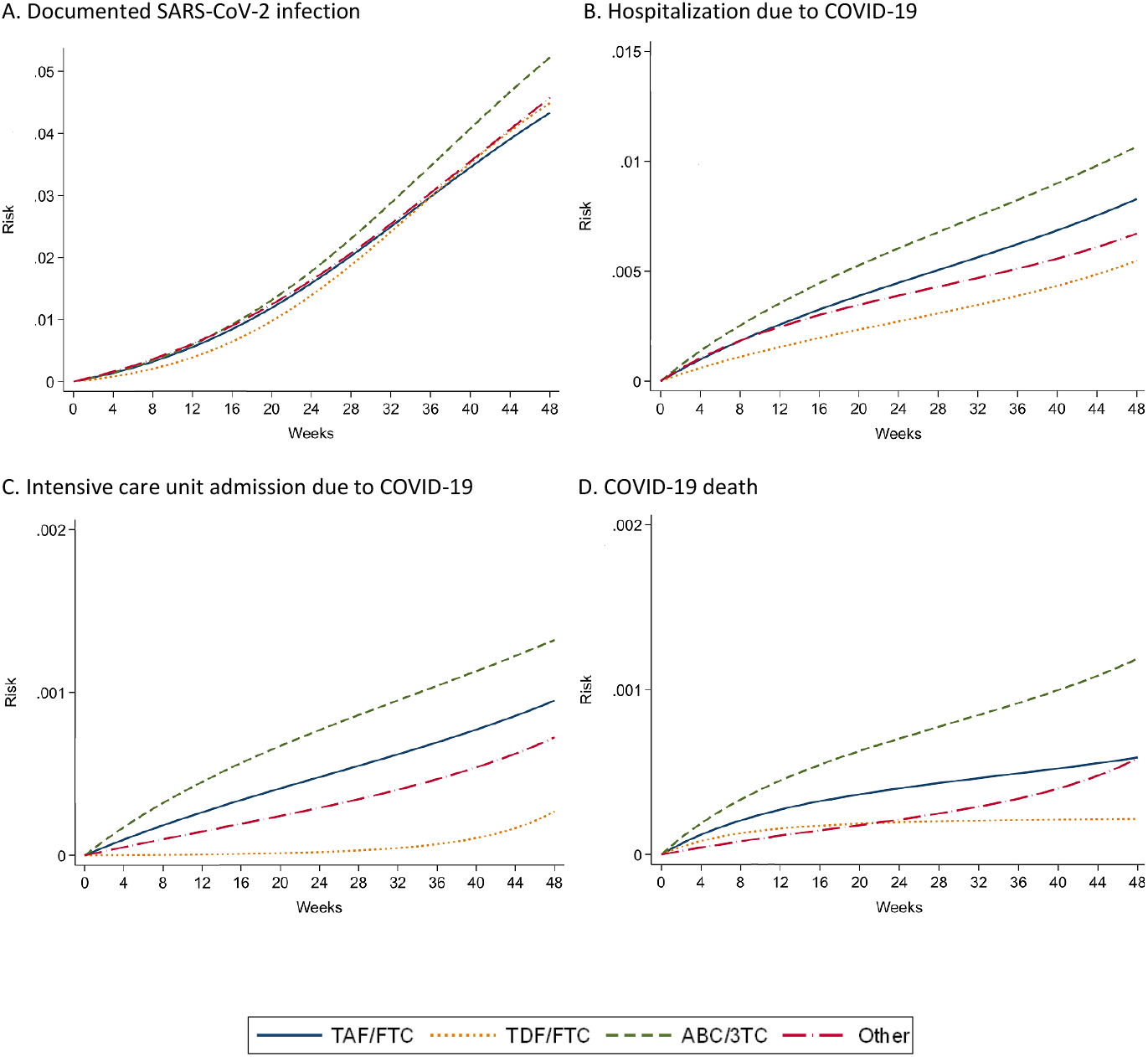
Estimated risks of COVID-19 outcomes by NRTI combination in HIV-positive individuals, ^*^CoVIHd Collaboration, Spain, February-December 2020 ^*^Adjusted via inverse probability weighting for age (in years, linear and quadratic terms), sex (male, female), transmission category (heterosexual, homo/bisexual, injecting drug use, other), country of origin (Spain, other), CD4 (<350, 350-500, >500 cells/mm3), and hypertension, diabetes, chronic renal disease, cardiovascular disease, and treatment with immunosuppressants or corticosteroids.

Table 2 shows the estimated 48-week risks of each outcome by NRTI combination. The estimated risk (95% CI) of documented SARS-CoV-2 infection was 4.3 % (4.1, 4.6) for TAF/FTC, 4.5% (3.9, 5.0) for TDF/FTC, and 5.2% (4.8, 5.6) for ABC/3TC. The estimated risks of COVID-19 hospitalization, ICU and death were lowest for TDF/FTC and highest for ABC/3TC (Table 2).

**Table 2.** Estimated 48-week risks, risk differences and risk ratios of COVID-19 outcomes by NRTI combination in HIV-positive individuals,* CoVIHd Collaboration, Spain, February-December 2020

|  | Documented SARS-CoV-2 infection |  |  |  | Hospitalization due to COVID-19 |  |  |  | ICU admission due to COVID-19 |  |  |  | COVID-19 death |  |  |  |
| --- | --- | --- | --- | --- | --- | --- | --- | --- | --- | --- | --- | --- | --- | --- | --- | --- |
|  | No. events | Risks (95%CI), % | Risk Differences (95% CI), % | Risk Ratios (95% CI) | No. events | Risks (95%CI), % | Risk Differences (95% CI), % | Risk Ratios (95% CI) | No. events | Risks (95% CI), % | Risk Differences (95% CI), % | Risk Ratios (95% CI) | No. events | Risks (95% CI), % | Risk Differences (95% CI), % | Risk Ratios (95% CI) |
| **TAF/FTC** | 923 | 4.3<br>(4.1, 4.6) | 0 | 1.00 | 157 | 0.8<br>(0.7, 1.0) | 0 | 1.00 | 17 | 0.09<br>(0.05,0.14) | 0 | 1.00 | 9 | 0.06<br>(0.02, 0.11) | 0 | 1.00 |
| **TDF/FTC** | 300 | 4.5<br>(3.9, 5.0) | 0.16<br>(-0.48, 0.69) | 1.04<br>(0.89, 1.17) | 35 | 0.5<br>(0.4, 0.7) | -0.28<br>(-0.52, -0.08) | 0.66<br>(0.43, 0.91) | 2 | 0.03<br>(0.01,0.08) | -0.07<br>(-0.12,-0.004) | 0.28<br>(0.11, 0.90) | 1 | 0.02<br>(0.02, 0.09) | -0.04<br>(-0.07, 0.03) | 0.37<br>(0.23, 1.90) |
| **ABC/3TC** | 687 | 5.2<br>(4.8, 5.6) | 0.89<br>(0.40, 1.34) | 1.21<br>(1.09, 1.33) | 147 | 1.1<br>(0.9, 1.2) | 0.24<br>(0.02, 0.45) | 1.29<br>(1.02, 1.58) | 18 | 0.13<br>(0.08,0.19) | 0.04<br>(-0.03,0.12) | 1.39<br>(0.70, 2.80) | 18 | 0.12<br>(0.07, 0.18) | 0.06<br>(-0.01, 0.12) | 2.02<br>(0.88, 6.12) |
| **Other regimes** | 492 | 4.6<br>(4.1, 5.0) | 0.24<br>(-0.27, 0.77) | 1.06<br>(0.94, 1.18) | 86 | 0.7<br>(0.5, 0.8) | -0.16<br>(-0.34,0.04) | 0.81<br>(0.62, 1.05) | 8 | 0.07<br>(0.02,0.12) | -0.02<br>(-0.09,0.04) | 0.76<br>(0.23, 1.77) | 9 | 0.06<br>(0.02, 0.11) | 0<br>(-0.06, 0.06) | 0.99<br>(0.34, 2.61) |

\* Adjusted via inverse probability weighting for age (in years, linear and quadratic terms), sex (male, female), transmission category (heterosexual, homo/bisexual, injecting drug use, other), country of origin (Spain, other), CD4 (<350, 350-500, >500 cells/mm<sup>3</sup>), and hypertension, diabetes, chronic renal disease, cardiovascular disease, and treatment with immunosuppressants or corticosteroids.

Compared with TAF/FTC, the estimated risk ratio (95% CI) of COVID-19 hospitalization was 0.66 (0.43, 0.91) for TDF/FTC, 1.29 (1.02, 1.58) for ABC/3TC, and 0.81 (0.62, 1.05) for others; the estimated risk ratio (95% CI) of COVID-19 ICU admission was 0.28 (0.11, 0.90) for TDF/FTC, 1.39 (0.70, 2.80) for ABC/3TC, and 0.76 (0.23, 1.77) for others; and the estimated risk ratio (95% CI) of COVID-19 death was 0.37 (0.23, 1.90) for TDF/FTC, 2.02 (0.88-6.12) for ABC/3TC, and 0.99 (0.34, 2.61) for others (Table 2).

Compared with TAF/FTC, the estimated risk ratios (95% CI) of asymptomatic SARS-CoV-2 infection and mild COVID-19 were greater than 1 for TDF/FTC and ABC/3TC. (Appendix Table 2).

Compared with TAF/FTC, the 48-week risk difference of hospitalizations per 1000 persons was -2.8 (95% CI -5.2 to -0.8) for TDF/FTC (Table 2). That is, the estimated number needed to treat with TDF/FTC vs. TAF/FTC during the study period would be 357 (192 to 1250) to prevent one hospitalization.

The estimates were similar in sensitivity analyses that censored at treatment switching (Appendix Table 3), that adjusted for confounding via standardization (Appendix Table 4) or a Cox model (Appendix Table 5), and that did not adjust for any covariates (Appendix Table 6).

The risk of COVID-19 hospitalization was similar across the three classes of third drug (Appendix Table 7).

Compared with TAF/FTC, the estimated risk ratio (95% CI) of COVID-19 hospitalization for TDF/FTC was 0.49 (0.24, 0.81) in individuals aged ≥50 years and 1.15 (0.59, 1.93) in younger individuals (Table 3). The corresponding risk ratio was similar in men and women (Appendix Table 8).

**Table 3.** Estimated 48-week risk, risk differences and risk ratios of COVID-19 outcomes by NRTI combination in HIV-positive individuals, stratified by age group, *CoVIHd Collaboration, Spain, February-December 2020

|  | Documented SARS-CoV-2 infection |  |  |  | Hospitalization due to COVID-19 |  |  |  |
| --- | --- | --- | --- | --- | --- | --- | --- | --- |
|  | No. events | Risks (95% CI), % | Risk Differences (95%CI), % | Risk Ratios (95%CI) | No. events | Risks (95% CI), % | Risk Differences (95%CI), % | Risk Ratios (95%CI) |
| <50 years |  |  |  |  |  |  |  |  |
| **TAF/FTC** | 561 | 4.9 (4.5, 5.3) | 0 | 1.00 | 48 | 0.4 (0.3, 0.6) | 0 | 1.00 |
| **TDF/FTC** | 198 | 5.3 (4.6, 6.0) | 0.45 (-0.43, 1.25) | 1.09 (0.91, 1.27) | 18 | 0.5 (0.3, 0.8) | 0.06 (-0.21, 0.34) | 1.15 (0.59, 1.93) |
| **ABC/3TC** | 336 | 5.4 (4.8, 6.0) | 0.50 (-0.27, 1.21) | 1.10 (0.95, 1.26) | 29 | 0.5 (0.3, 0.6) | 0.02 (-0.22, 0.22) | 1.03 (0.59, 1.62) |
| **Other regimes** | 220 | 5.4 (4.7, 6.0) | 0.47 (-0.35, 1.23) | 1.10 (0.93, 1.26) | 17 | 0.4 (0.2, 0.6) | -0.04 (-0.26, 0.17) | 0.91 (0.49, 1.47) |
| ≥50 years |  |  |  |  |  |  |  |  |
| **TAF/FTC** | 362 | 3.8 (3.4, 4.2) | 0 | 1.00 | 109 | 1.2 (1.0, 1.4) | 0 | 1.00 |
| **TDF/FTC** | 102 | 3.6 (2.9, 4.3) | -0.17 (-0.94, 0.64) | 0.96 (0.77, 1.18) | 17 | 0.6 (0.3, 0.9) | -0.61 (-1.03, -0.22) | 0.49 (0.24, 0.81) |
| **ABC/3TC** | 351 | 5.0 (4.6, 5.6) | 1.26 (0.62, 1.89) | 1.33 (1.15, 1.55) | 118 | 1.7 (1.4, 2.0) | 0.47 (0.11, 0.83) | 1.40 (1.08, 1.79) |
| **Other regimes** | 272 | 3.9 (3.4, 4.3) | 0.07 (-0.51, 0.64) | 1.02 (0.87, 1.18) | 69 | 0.9 (0.7, 1.1) | -0.29 (-0.62, 0.03) | 0.76 (0.54, 1.03) |

\* Adjusted via inverse probability weighting for age (in years, linear and quadratic terms), sex (male, female), transmission category (heterosexual, homo/bisexual, injecting drug use, other), country of origin (Spain, other), CD4 (<350, 350-500, >500 cells/mm<sup>3</sup>), and hypertension, diabetes, chronic renal disease, cardiovascular disease, and treatment with immunosuppressants or corticosteroids.

Compared with all NRTI combinations without TDF, the estimated risk ratios (95% CI) of COVID-19 hospitalization, ICU admission and death were 0.64 (0.42-0.89), 0.28 (0.11-0.84) and 0.29 (0.20-1.11), respectively, for TDF/FTC. In individuals aged ≥50 years, these risk ratios were 0.48 (0.24-0.76), 0.24 (0.18-0.88) and 0.22 (0.15-0.97).

## Discussion

We studied over 50,000 HIV-positive individuals on antiretroviral therapy with adequate virological control in Spain during 2020, before the start of the SARS-CoV-2 vaccination campaign. The estimated risks of COVID-19 hospitalization and ICU admission were lower among individuals treated with TDF/FTC than among those treated with other antiretrovirals. The potential benefit of TDF/FTC appeared to be restricted to individuals over 50 years of age who have a higher risk of developing severe COVID-19. In this age group, the risk of COVID-19 hospitalization was about 50% lower for TDF/FTC compared with TAF/FTC, the most commonly used NRTI combination. The risk of death from COVID-19 was also lower for antiretroviral regimes based on TDF/FTC, but the estimates were very imprecise. The estimated risks of documented infection and of mild infection are difficult to interpret because of incomplete ascertainment.

Our estimates are consistent with those from observational studies conducted during the first epidemic wave in South Africa^6^ and Spain^5^. These studies, which preferentially included COVID-19 cases that were severe enough to be diagnosed during a period in which only a fraction of cases were confirmed, found a lower risk of severe COVID-19 among HIV-positive individuals on TDF/FTC compared with other NRTI combinations. The latter study did not collect information on comorbidities, used reported population frequencies of antiretroviral use for non-cases (which resulted in a slight overestimation of the proportion of the population on TDF/FTC and an underestimation of the proportion on TAF/FTC), and did not restrict the analyses to persons with virological suppression^5,6^. Another study in Spain found a lower SARS-CoV-2 prevalence among TDF/FTC users than in TAF/FTC users^7^. The present study improves upon previous ones because it simultaneously includes a large population of HIV-positive individuals with adequate antiretroviral control and adjusts for multiple comorbidities.

Importantly, adjustment for comorbidities and other factors had little impact on our effect estimates, a finding that was not unexpected in our setting^13^ and which we confirm in this study. Because the association between TDF/FTC and a lower risk of severe COVID-19 cannot be fully explained by differences between individuals receiving TDF/FTC and individuals receiving other antiretrovirals, it is likely that the observed association is the result of a causally protective effect of TDF/FTC in HIV-positive individuals. In contrast, individuals on ABC/3TC had a higher risk of severe COVID-19 than those on other NRTI combinations. While persons on ABC/3TC (and those on regimes based on dual therapy) are older and have a higher prevalence of comorbidities, the higher relative risk in this group persisted even after adjustment for these factors.

A protective effect of TDF/FTC is biologically plausible. *In silico* studies suggest that all forms of tenofovir, like other nucleos(t)ide analogues, partly inhibit the SARS-CoV-2 RNA-dependent RNA-polymerase (RNAdRNAp)^14–16^ and some, but not all, *in vitro* studies also suggest that tenofovir inhibits the RNAdRNAp^17,18^. Because of the possible higher bioavailability of TDF than TAF in respiratory cells, TDF might result in greater inhibition of the SARS-CoV-2 RNApRNAp^19–23^. In addition, tenofovir has been reported to have immunomodulatory effects^24–27^ and animal models suggest that TDF/FTC increases nasopharyngeal SARS-CoV-2 clearance^28^. The possibility of TDF-mediated viral inhibition is further supported by a recent randomized trial that found that TDF/FTC accelerates nasopharyngeal SARS-CoV-2 clearance in HIV-negative individuals within 7 days of COVID-19 symptoms^29^ and another one (not yet peer-reviewed) that reported a lower risk of invasive mechanical ventilation in patients on TDF/FTC + colchicine + rosuvastatin combination^30^. Three ongoing randomized trials are studying the effects of TDF/FTC on clinical outcomes: The EPICOS trial studies TDF/FTC as pre-exposure prophylaxis among healthcare workers in Spain and Latin America^31^, the PANCOVID trial studies TDF/FTC as treatment of COVID-19 in Spain^32^ and the ARTAN-C19 trial in Brazil^33^. The two Spanish trials are led by some of the co-authors of this report.

While the COVID-19 pandemic continues, a protective effect of TDF/FTC has clinical implications for HIV-positive individuals because TDF/FTC is an effective drug to control HIV infection in individuals without impaired renal function^34,35^. Similar considerations apply to individuals with hepatitis B infection. It is possible that a similar protection could extend to HIV-negative individuals, which would be especially important for immunosuppressed patients for whom vaccines have suboptimal effectiveness. Compared with some other drugs repurposed for COVID-19, TDF has several advantages. First, it has a solid safety track record in individuals with normal renal function^34^, including pregnant women^35,36^, and in fact is used routinely as pre-exposure prophylaxis for HIV infection. Second, it is administered orally and thus does not need to be administered in a healthcare facility. Third, it is an inexpensive generic drug that could be massively produced in many countries, including in settings with low COVID-19 vaccine coverage.

Our study has some limitations. First, residual confounding by yet to be identified factors cannot be excluded. However, such residual confounding seems unlikely because we adjusted for known factors that affect both antiretroviral treatment choice and COVID-19 severity. Therefore, the lowest risk of hospitalization in those receiving TDF/FTC cannot be easily explained by residual confounding. Second, we could not collect SARS-CoV-2 testing frequency for all individuals. Testing patterns, however, are expected to affect the detection of mild (and asymptomatic) disease but not that of more severe outcomes like hospitalization, ICU admissions, and death. Third, missing data on comorbidities led to the exclusion of 22% of otherwise eligible individuals. However, estimates did not materially change in unadjusted analyses that included individuals with missing data on comorbidities. Fourth, even a large cohort like this one cannot provide precise estimates for the risks of infrequent events such as ICU admissions and deaths.

In summary, our findings suggest that treatment with TDF/FTC results in a lower severity of COVID-19 than treatment with other antiretrovirals among HIV-positive individuals with adequate virological control. This protective effect may be largely restricted to individuals aged 50 years and older. Confirmatory randomized trials of TDF/FTC for the prophylaxis and early treatment of COVID-19 are warranted.

## Data Availability

All data produced in the present study are available upon reasonable request to the authors

