## Appendix for "Tenofovir Disoproxil Fumarate and severity of COVID-19 in people with HIV infection"

Appendix Table 1. Description of antiretroviral regimes in each NRTI combination in HIV-positive individuals, CoVIHd Collaboration, Spain, February-December 2020

| TAF/FTC **N = 20,432** | | TDF/FTC **N = 6,160** | | ABC/3TC **N = 13,715** | | Other **N = 11,251** | |
|---|---|---|---|---|---|---|---|
| **BIC/TAF/FTC** | 7,517 (37.1) | EFV/TDF/FTC | 2,186 (35.5) | DTG/ABC/3TC | 9,204 (67.1) | DTG/3TC | 3,832 (34.1) |
| **EVG/COBI/TAF/FTC** | 3,937 (19.4) | RPV/TDF/FTC | 1,340 (21.7) | NVP+ABC/3TC | 1,083 (7.9) | DTG/RPV | 1,747 (15.5) |
| **RPV/TAF/FTC** | 3,695 (18.2) | RAL+TDF/FTC | 692 (11.2) | RPV+ABC/3TC | 1,043 (7.6) | bDRV | 1,688 (15.0) |
| **DRV/COBI/TAF/FTC** | 3,210 (15.8) | bDRV+TDF/FTC | 624 (10.1) | RAL+ABC/3TC | 961 (7.0) | bDRV+3TC | 1,200 (10.7) |
| **RAL+TAF/FTC** | 744 (3.7) | DTG+TDF/FTC | 619 (10.0) | bDRV+ABC/3TC | 560 (4.1) | bDRV+DTG | 796 (7.1) |
| **DTG+TAF/FTC** | 682 (3.4) | NVP+TDF/FTC | 325 (5.3) | EFV+ABC/3TC | 554 (4.0) | bDRV+RAL | 520 (4.6) |
| **NVP+TAF/FTC** | 396 (1.9) | EVG/COBI/TDF/FTC | 140 (2.3) | ATVr+ABC/3TC | 108 (0.8) | bDRV+RPV | 221 (2.0) |
| **EFV+TAF/FTC** | 91 (0.4) | bATV+TDF/FTC | 86 (1.4) | ETV+ABC/3TC | 87 (0.6) | bDRV+ETV | 144 (1.3) |
| **Other** | 160 (0.8) | LPVr+TDF/FTC | 50 (0.8) | Other | 115 (0.8) | RAL+ETV | 143 (1.3) |
| | | ETV+TDF/FTC | 42 (0.7) | | | DTG | 136 (1.2) |
| | | Other | 56 (0.9) | | | CAB+RPV | 126 (1.1) |
| | | | | | | RAL+3TC | 112 (1.0) |
| | | | | | | Other | 586 (5,2) |

ABC, abacavir; BIC, Bictegravir; bATV, Atazanavir boosted; bDRV, Darunavir boosted; CAB, Cabotegravir; COBI, Cobicistat; DRV, Darunavir; DTG, Dolutegravir; EFV, Efavirenz; EVG, Elvitegravir; ETV, Etravirine; FTC, Emtricitabine; LPVr, Lopinavir/ritonavir; RAL, RaltegravirNVP, Nevirapine; RPV, Rilpivirine; TAF, Tenofovir alafenamide; 3TC, Lamivudine.

Appendix Table 2. Estimated 48-week risks, risk differences and risk ratios of asymptomatic SARS-CoV-2 infection and mild COVID-19 by NRTI combination in HIV-positive individuals,* CoVIHd Collaboration, Spain, February-December 2020

| | Asymptomatic SARS-CoV-2 infection | | | | Mild COVID-19 | | | |
|---|---|---|---|---|---|---|---|---|
| | **No. events** | **Risks (95% CI), %** | **Risk Differences (95%CI), %** | **Risk Ratios (95%CI)** | **No. events** | **Risks (95% CI), %** | **Risk Differences (95%CI), %** | **Risk Ratios (95%CI)** |
| **TAF/FTC** | 206 | 1.00 (0.87, 1.12) | 0 | 1.00 | 389 | 1.82 (1.64, 2.01) | 0 | 1.00 |
| **TDF/FTC** | 77 | 1.15 (0.91, 1.45) | 0.15 (-0.13, 0.44) | 1.15 (0.87, 1.46) | 155 | 2.21 (1.82, 2.58) | 0.39 (-0.07, 0.76) | 1.21 (0.97, 1.43) |
| **ABC/3TC** | 133 | 1.06 (0.89, 1.26) | 0.07 (-0.16, 0.29) | 1.07 (0.85, 1.31) | 274 | 2.16 (1.91, 2.42) | 0.34 (0.01, 0.65) | 1.19 (1.00, 1.38) |
| **Other regimes** | 123 | 1.16 (0.96, 1.38) | 0.17 (-0.06, 0.41) | 1.17 (0.95, 1.44) | 219 | 2.10 (1.84, 2.42) | 0.28 (-0.04, 0.64) | 1.15 (0.98, 1.37) |

* Adjusted via inverse probability weighting for age (in years, linear and quadratic terms), sex (male, female), transmission category (heterosexual, homo/bisexual, injecting drug use, other), country of origin (Spain, other), CD4 (<350, 350-500, >500 cells/mm3), and hypertension, diabetes, chronic renal disease, cardiovascular disease, and treatment with immunosuppressants or corticosteroids.

Appendix Table 3. Per-protocol analysis: Estimated 48-week risk differences and risk ratios of COVID-19 outcomes by NRTI combination in HIV-positive individuals,* CoVIHd Collaboration, Spain, February-December 2020

| | **Documented SARS-CoV-2 infection** | | **Hospitalization due to COVID-19** | | **ICU admission due to COVID-19** | | **COVID-19 death** | |
|---|---|---|---|---|---|---|---|---|
| | **Risk Differences (95% CI), %** | **Risk Ratios (95% CI)** | **Risk Differences (95% CI), %** | **Risk Ratios (95% CI)** | **Risk Differences (95% CI), %** | **Risk Ratios (95% CI)** | **Risk Differences (95% CI), %** | **Risk Ratios (95% CI)** |
| **TAF/FTC** | 0 | 1.00 | 0 | 1.00 | 0 | 1.00 | 0 | 1.00 |
| **TDF/FTC** | 0.10 (-0.57, 0.68) | 1.02 (0.88, 1.16) | -0.27 (-0.54, -0.04) | 0.68 (0.43, 0.95) | -0.06 (-0.12, 0.01) | 0.33 (0.12, 1.07) | -0.04 (-0.08, 0.03) | 0.37 (0.23, 1.93) |
| **ABC/3TC** | 0.87 (0.36, 1.37) | 1.19 (1.08, 1.32) | 0.22 (-0.003, 0.45) | 1.26 (1.00, 1.58) | 0.04 (-0.04, 0.13) | 1.44 (0.64, 2.94) | 0.04 (-0.03, 0.10) | 1.61 (0.68, 4.44) |
| **Other regimes** | 0.56 (0.04, 1.12) | 1.13 (1.01, 1.26) | -0.13 (-0.33, 0.07) | 0.85 (0.64, 1.10) | -0.01 (-0.09, 0.06) | 0.86 (0.25, 2.08) | 0.004 (-0.06,0.07) | 1.07 (0.36, 2.75) |

* Adjusted via inverse probability weighting for age (in years, linear and quadratic terms), sex (male, female), transmission category (heterosexual, homo/bisexual, injecting drug use, other), country of origin (Spain, other), CD4 (<350, 350-500, >500 cells/mm3), and hypertension, diabetes, chronic renal disease, cardiovascular disease, and treatment with immunosuppressants or corticosteroids.

Appendix Table 4. Standardized 48-week risk differences and risk ratios of COVID-19 outcomes by NRTI combination in HIV-positive individuals,* CoVIHd Collaboration, Spain, February-December 2020

| | **Documented SARS-CoV-2 infection** | | **Hospitalization due to COVID-19** | | **ICU admission due to COVID-19** | | **COVID-19 death** | |
|---|---|---|---|---|---|---|---|---|
| | **Risk Differences (95% CI), %** | **Risk Ratios (95% CI)** | **Risk Differences (95% CI), %** | **Risk Ratios (95% CI)** | **Risk Differences (95% CI), %** | **Risk Ratios (95% CI)** | **Risk Differences (95% CI), %** | **Risk Ratios (95% CI)** |
| **TAF/FTC** | 0 | 1.00 | 0 | 1.00 | 0 | 1.00 | 0 | 1.00 |
| **TDF/FTC** | 0.34 (-0.28, 0.89) | 1.08 (0.94, 1.21) | -0.25 (-0.52, 0.003) | 0.73 (0.47, 1.00) | -0.06 (-0.12, 0.02) | 0.40 (0.15, 1.26) | -0.03 (-0.06, 0.04) | 0.45 (0.28, 2.14) |
| **ABC/3TC** | 0.87 (0.40, 1.32) | 1.20 (1.09, 1.32) | 0.28 (0.03, 0.49) | 1.30 (1.03, 1.61) | 0.05 (-0.03, 0.14) | 1.51 (0.77, 3.04) | 0.07 (0.01, 0.13) | 2.23 (1.10, 5.77) |
| **Other regimes** | 0.21 (-0.29, 0.72) | 1.05 (0.93, 1.17) | -0.15 (-0.35, 0.06) | 0.83 (0.64, 1.07) | -0.02 (-0.10, 0.05) | 0.75 (0.24, 1.75) | 0.003 (-0.04,0.06) | 1.06 (0.40, 2.65) |

* Adjusted via standardization for age (in years, linear and quadratic terms), sex (male, female), transmission category (heterosexual, homo/bisexual, injecting drug use, other), country of origin (Spain, other), CD4 (<350, 350-500, >500 cells/mm3), and hypertension, diabetes, chronic renal disease, cardiovascular disease, and treatment with immunosuppressants or corticosteroids.

Appendix Table 5. Estimated hazard ratios of COVID-19 outcomes by NRTI combination in HIV-positive individuals,* CoVIHd Collaboration, Spain, February-December 2020

| | Documented SARS-CoV-2 infection | Hospitalization due to COVID-19 | ICU admission due to COVID-19 | COVID-19 death |
|---|---|---|---|---|
| | HR (95% CI) | | | |
| **TAF/FTC** | 1.00 | 1.00 | 1.00 | 1.00 |
| **TDF/FTC** | 1.08 (0.93, 1.21) | 0.73 (0.47, 1.00) | 0.40 (0, 1.26) | 0.44 (0, 1.98) |
| **ABC/3TC** | 1.21 (1.09, 1.33) | 1.31 (1.03, 1.62) | 1.51 (0.77, 3.18) | 2.25 (1.09, 5.60) |
| **Other regimes** | 1.05 (0.93, 1.18) | 0.83 (0.63, 1.07) | 0.75 (0.24, 1.83) | 1.06 (0.41, 2.69) |

* Adjusted from Cox model with covariates age (in years, linear and quadratic terms), sex (male, female), transmission category (heterosexual, homo/bisexual, injecting drug use, other), country of origin (Spain, other), CD4 (<350, 350-500, >500 cells/mm3), and hypertension, diabetes, chronic renal disease, cardiovascular disease, and treatment with immunosuppressants or corticosteroids.

Appendix Table 6. Unadjusted 48-week risk differences and risk ratios of COVID-19 outcomes by NRTI combination in HIV-positive individuals, CoVIHd Collaboration, Spain, February-December 2020

| | Documented SARS-CoV-2 infection | | Hospitalization due to COVID-19 | | ICU admission due to COVID-19 | | COVID-19 death | |
|---|---|---|---|---|---|---|---|---|
| | **Risk Differences (95% CI), %** | **Risk Ratios (95% CI)** | **Risk Differences (95% CI), %** | **Risk Ratios (95% CI)** | **Risk Differences (95% CI), %** | **Risk Ratios (95% CI)** | **Risk Differences (95% CI), %** | **Risk Ratios (95% CI)** |
| **TAF/FTC** | 0 | 1.00 | 0 | 1.00 | 0 | 1.00 | 0 | 1.00 |
| **TDF/FTC** | 0.35 (-0.29, 0.92) | 1.08 (0.94, 1.21) | -0.20 (-0.44, 0.01) | 0.74 (0.48, 1.01) | -0.05 (-0.10, 0.01) | 0.39 (0.14, 1.21) | -0.03 (-0.06, 0.02) | 0.37 (0.22, 1.71) |
| **ABC/3TC** | 0.49 (0.02, 0.95) | 1.11 (1.00, 1.22) | 0.30 (0.07, 0.50) | 1.39 (1.09, 1.73) | 0.05 (-0.02, 0.12) | 1.58 (0.83, 3.16) | 0.09 (0.03, 0.15) | 2.98 (1.48, 8.62) |
| **Other regimes** | -0.14 (-0.64, 0.37) | 0.97 (0.86, 1.08) | -0.0.004 (-0.20, 0.18) | 0.99 (0.77, 1.28) | -0.01 (-0.07, 0.05) | 0.85 (0.29, 1.81) | 0.04 (-0.02, 0.10) | 1.82 (0.66, 4.58) |

Appendix Table 7. Estimated 48-week risks, risk differences and risk ratios of COVID-19 outcomes by third antiretroviral drug in HIV-positive individuals,* CoVIHd Collaboration, Spain, February-December 2020

| | Documented SARS-CoV-2 infection | | | | Hospitalization due to COVID-19 | | | | ICU admission due to COVID-19 | | | | COVID-19 death | | | |
|---|---|---|---|---|---|---|---|---|---|---|---|---|---|---|---|---|
| | **No. events** | **Risks (95%CI), %** | **Risk Differences (95% CI), %** | **Risk Ratios (95% CI)** | **No. events** | **Risks (95%CI), %** | **Risk Differences (95% CI), %** | **Risk Ratios (95% CI)** | **No. events** | **Risks (95% CI), %** | **Risk Differences (95% CI), %** | **Risk Ratios (95% CI)** | **No. events** | **Risks (95% CI), %** | **Risk Differences (95% CI), %** | **Risk Ratios (95% CI)** |
| **II** | 1,140 | 4.6 (4.3, 4.9) | 0 | 1.00 | 214 | 0.8 (0.7, 1.0) | 0 | 1.00 | 23 | 0.08 (0.05,0.12) | 0 | 1.00 | 12 | 0.04 (0.02,0.07) | 0 | 1.00 |
| **PI** | 231 | 5.0 (4.2, 5.7) | 0.35 (-0.48,1.19) | 1.08 (0.90, 1.27) | 42 | 1.0 (0.6, 1.4) | 0.12 (-0.28,0.56) | 1.14 (0.70, 1.71) | 4 | 0.08 (0.02,0.17) | -0.007 (-0.07,0.08) | 0.92 (0.20, 2.22) | 4 | 0.15 (0.03,0.36) | 0.11 (-0.02,0.31) | 3.69 (0.62,10.77) |
| **NNRTI** | 533 | 5.2 (4.7, 5.7) | 0.62 (0.03, 1.21) | 1.13 (1.01, 1.28) | 83 | 0.9 (0.7, 1.1) | 0.04 (-0.20,0.29) | 1.04 (0.77, 1.37) | 10 | 0.16 (0.06,0.30) | 0.08 (-0.02,0.22) | 2.00 (0.74, 4.25) | 12 | 0.19 (0.09,0.32) | 0.15 (0.04, 0.28) | 4.47 (1.68,11.73) |

II: Integrase inhibitor, PI: protease inhibitor, NNRTI: non-nucleoside reverse-transcriptase inhibitor

* Adjusted via inverse probability weighting for NRTI combination (TAF/FTC, TDF/FTC, ABC/3TC), age (in years, linear and quadratic terms), sex (male, female), transmission category (heterosexual, homo/bisexual, injecting drug use, other), country of origin (Spain, other), CD4 (<350, 350-500, >500 cells/mm3), and hypertension, diabetes, chronic renal disease, cardiovascular disease, and treatment with immunosuppressants or corticosteroids.

Appendix Table 8. Estimated 48-week risks, risk differences and risk ratios of COVID-19 outcomes by NRTI combination in HIV-positive individuals, stratified by sex,* CoVIHd Collaboration, Spain, February-December 2020

| | Documented SARS-CoV-2 infection | | | | Hospitalization due to COVID-19 | | | |
|---|---|---|---|---|---|---|---|---|
| | **No. events** | **Risks (95% CI), %** | **Risk Differences (95%CI), %** | **Risk Ratios (95%CI)** | **No. events** | **Risks (95% CI), %** | **Risk Differences (95%CI), %** | **Risk Ratios (95%CI)** |
| Men | | | | | | | | |
| **TAF/FTC** | 734 | 4.3 (4.0, 4.6) | 0 | 1.00 | 126 | 0.8 (0.7, 1.0) | 0 | 1.00 |
| **TDF/FTC** | 228 | 4.3 (3.8, 4.9) | 0.08 (-0.51, 0.72) | 1.02 (0.88, 1.18) | 26 | 0.5 (0.3, 0.8) | -0.29 (-0.56, 0.01) | 0.64 (0.40, 1.01) |
| **ABC/3TC** | 554 | 5.3 (4.9, 5.8) | 1.08 (0.52, 1.60) | 1.25 (1.12, 1.39) | 123 | 1.1 (0.9, 1.3) | 0.30 (0.06, 0.55) | 1.36 (1.06, 1.75) |
| **Other regimes** | 385 | 4.7 (4.2, 5.1) | 0.40 (-0.15, 0.99) | 1.09 (0.97, 1.24) | 71 | 0.7 (0.6, 0.9) | -0.11 (-0.32, 0.13) | 0.86 (0.64, 1.18) |
| Women | | | | | | | | |
| **TAF/FTC** | 189 | 4.6 (4.0, 5.3) | 0 | 1.00 | 31 | 0.8 (0.6, 1.2) | 0 | 1.00 |
| **TDF/FTC** | 72 | 5.0 (3.9, 6.2) | 0.45 (-0.86, 1.68) | 1.10 (0.83, 1.41) | 9 | 0.6 (0.2, 1.1) | -0.23 (-0.70, 0.27) | 0.73 (0.27, 1.45) |
| **ABC/3TC** | 133 | 4.8 (3.9, 5.6) | 0.19 (-0.83, 1.11) | 1.04 (0.83, 1.26) | 24 | 0.8 (0.5, 1.2) | 0.005 (-0.44, 0.40) | 1.01 (0.58, 1.61) |
| **Other regimes** | 107 | 4.2 (3.5, 5.0) | -0.33 (-1.34, 0.66) | 0.93 (0.72, 1.16) | 15 | 0.5 (0.3, 0.8) | -0.33 (-0.72, 0.06) | 0.61 (0.28, 1.10) |

* Adjusted via inverse probability weighting for age (in years, linear and quadratic terms), transmission category (heterosexual, homo/bisexual, injecting drug use, other), country of origin (Spain, other), CD4 (<350, 350-500, >500 cells/mm3), and hypertension, diabetes, chronic renal disease, cardiovascular disease, and treatment with immunosuppressants or corticosteroids.

Appendix Figure 1. Estimated risks of asymptomatic SARS-CoV-2 infection and mild COVID-19 by NRTI combination in HIV-positive individuals,* CoVIHd Collaboration, Spain, February-December 2020

A. Asymptomatic SARS-CoV-2 infection

B. Mild COVID-19

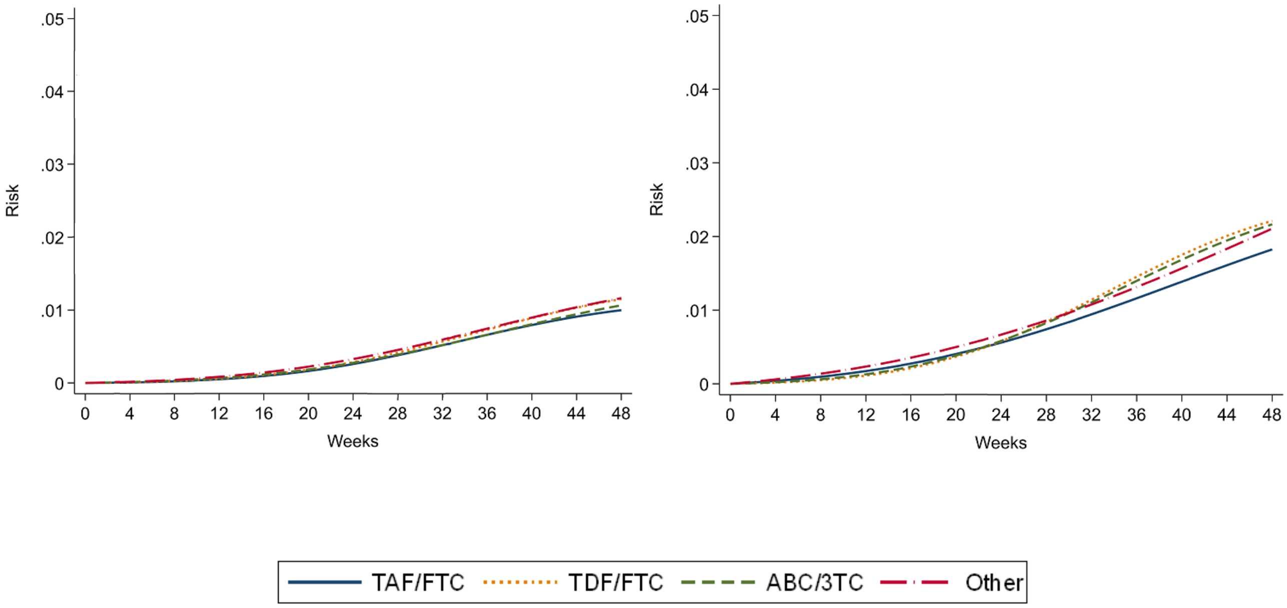


* Adjusted via inverse probability weighting for age (in years, linear and quadratic terms), sex (male, female), transmission category (heterosexual, homo/bisexual, injecting drug use, other), country of origin (Spain, other), CD4 (<350, 350-500, >500 cells/mm3), and hypertension, diabetes, chronic renal disease, cardiovascular disease, and treatment with immunosuppressants or corticosteroids.

**Appendix 1: Investigators of the CoVIHd Collaboration in Spain**

*Principal investigators (PI) and co-investigators of the participating hospitals by Spanish region*

Andalucía

Hospital Virgen del Rocío: PIs: LF. López Cortés, N. Espinosa. Co-investigators: S. Llaves, C. Roca, M.Herrero, C.Sotomayor, MJ.Rodriguez Hernandez, JM.Cisneros.

Hospital Universitario Virgen de la Victoria: PIs: J. Santos, R.Palacios. Co-investigators: C. Gómez-Ayerve, I.Pérez-Hernández, A.Prolo, M.Villalobos, M.López-Jódar, I.Viciana.

Hospital Costa del Sol: PIs: J. Olalla, A del Arco Jiménez. Co-investigators: J Pérez Stachowski, JM García de Lomas, MD Martín Escalante, J García Alegría, MA Onieva, F Fernández Sánchez.

Hospital Virgen de las Nieves: PIs: C. Hidalgo Tenorio, J.Pasquau. Co-investigators: C. García, S. Sequera.

Hospital Universitario de Valme: PIs: J. Macías. Co-investigators: P. Rincón, M. Fernández.

Hospital Universitario Juan Ramón Jiménez: PIs: D. Merino,¡. Co-investigators: L. Corpa, M. Raffo, A.Jimenez, M.Franco,

Hospital Regional Universitario de Málaga: PIs: M.Castaño, M. Delgado.

Hospital Universitario Reina Sofía: PIs: A. Rivero, A. Rivero-Juarez.

Hospital Universitario Torrecárdenas: PIs: A. Collado.

Hospital Universitario de Puerto Real: PIs: A. Romero.

Aragón

Hospital Clínico Universitario Lozano Blesa: PIs: I. Sanjoaquín. Co-investigators: M. Gimeno, JM. Vinuesa, S. Letona, MJ. Crusells.

Hospital General San Jorge: PIs: M. Egido. Co-investigators: T. Omiste.

Hospital Miguel Servet: PIs: P. Arazo, R. García.

Canarias

Hospital Insular de Las Palmas: PIs: JL. Pérez-Arellano, C. Lavilla. Co-investigators: L. Suárez-Hormiga, L. López-Delgado, A. Granados, M. Hernandez-Cabrera, E. Pisos, N. Jaén.

Hospital Universitario de Canarias: PIs: JL Gómez Sirvent, MR.Alemán. Co-investigators: MM.Alonso, R.Pelazas, D.García Rosado, AM.López-Lirola, I. Hernández.

Cantabria

Hospital Universitario Marqués de Valdecilla: PIs: MC. Fariñas, F. Arnaiz de las Revillas

Co-investigators: C. González-Rico, A. Illaro; J. Calvo, ME. Cano, M. Gutierrez- Cuadra, C. Armiñanzas.

Castilla y León

Hospital Universitario de Burgos: PIs: C. Navarro-San Francisco. Co-investigators: M Fernandez Regueras

Hospital Clínico Universitario de Valladolid: PIs: C. Dueñas. Co-investigators: E. Tapia, S. Gutiérrez, G. Zapico, L. Rodríguez.

Hospital Río Hortega de Valladolid: PIs: J. Gómez. Co-investigators: M.Cobos, M. González, AM. Corcho, J. Abadía.

Hospital Universitario de León: PIs: JM. Guerra.

Complejo Asistencial Universitario de Palencia: PIs: JJ. Sánchez. Co-investigators: C. Sanchez, Y. Morán.

Complejo Asistencial Ntra. Sra. de Sonsoles de Ávila: PIs: MA. Garcinuño. Co-investigators: C. Grande, AC. Antolí, M.Pedromingo,JM.Barragán-Casas.

Hospital General de Segovia: PIs: EM.Ferreira. Co-investigators: P.Bachiller, AM. Carrero, S.Muñóz, JM Alonso de los Santos.

Castilla La Mancha

Complejo Hospitalario de Toledo: PIs: F. Cuadra. Co-investigators: J. Largo, MA. Sepulveda.

Hospital General La Mancha Centro: PIs: JR. Barberá. Co-investigators: E. Arroyo.

Hospital Virgen de la Luz: PIs: P. Geijo

Hospital General Universitario de Albacete: PIs: E. Martínez.

Cataluña

Hospital Clínic Barcelona: PIs E. Martínez, JL. Blanco, E. de Lazzari. Co-investigators: JM.Miró, J.Mallolas, M.Laguno, M.Martínez-Rebollar, B.Torres, A.Iniciarte, A.González Cordón, L. de la Mora.

Hospital Universitari Vall d'Hebron: PIs: A. Curran, V. Falcó. Co-investigators: JN. García, J. Burgos, J. Navarro, P. Suanzes, M. Sanchiz, I. Rodriguez.

Hospital Universitari de Bellvitge: PIs: A. Imaz, D. Podzamczer. Co-investigators: S. Scévola, P. Prieto, A. Silva, M. Saumoy, J. Tiraboschi.

Hospital Santa María: PIs E. González. Co-investigators: N. Abdulghani, R. Sola, T. Comella, L. Gutierrez.

Hospital Universitari Mútua Terrassa: PIs: D. Dalmau. Co-investigators: M. Cairó, M. Martinez, R. Font, X.Martínez-Lacasa

Consorci Corporació Sanitària Parc Taulí de Sabadell: PIs G. Navarro. Co-investigators: S. Calzado, M. Navarro.

Parc Sanitari Sant Joan de Déu: PIs: V. Diaz-Brito. Co-investigators: A. Delicado, M. Sanmartí, E. Moreno, F. Medina.

Hospital Universitari Joan XXIII de Tarragona: PIs: F. Vidal. Co-investigators: E. Yeregui, A. Marti, A. Rull, J. Peraire

Consorcio Sanitario del Maresme (Hospital de Mataró): PIs L. Force. Co-investigators: P. Barrufet, L.Arbones, L.Albiach.

Hospital Dr. Josep Trueta: PIs A. Oller. Co-investigators: X. Salgado, M. Lora.

Consorcio Hospitalario de Vic: PIs I. Vilaró. Co-investigators: MJ. Martínez.

Consorci Sanitari de Terrasa:PIs M. Aranda. Co-investigators: R. Solé, M. Roca.

Hospital de Viladecans: PIs A. Lérida. Co-investigators: M. Muelas, A. Figueras, L. Escolano, N.Carrasco Fons.

Hospital Sant Jaume de Calella: PIs O. del Río. Co-investigators: S. Valero.

Hospital de Mollet: PIs P. Vázquez. Co-investigators: JM. Tricas, M. Priegue.

Fundació Salut Empordà (Figueres): PIs J. Cucurull. Co-investigators: S. Vega, JA. Damián.

Hospital de Palamós: PIs: M.Hortos. Co-investigators: A. Masabeu

Hospital de Tortosa Verge de la Cinta: PIs: A. Orti. Co-investigators: E. Chamarro, C.Escrig

Hospital General de Granollers: PIs: E. Deig.

Comunidad de Madrid

Hospital Universitario LA PAZ- H. Carlos III: PIs: JR. Arribas, R.Montejano, J.Cadiñanos.
Co-investigators: C. Busca, R. Micán, R.de Miguel-Buckley, JI.Bernardino, M.Montes, J.González-García, L.Ramos, L.Martín-Carbonero.

Hospital Fundación Jiménez Díaz: PIs: A. Cabello, M.Górgolas, A.Herrero. Co-investigators: B.Álvarez Álvarez, L.Prieto Pérez, I.Carrillo Acosta, A.Al-Hayani, R.Fernández Doblas, R.Téllez, S.Heili-Frades, JM. Milicua.

Hospital Universitario Ramón y Cajal: PIs: S. Moreno, MJ. Pérez, S. del Campo. Co-investigators:, JA. Pérez, P. Vizcarra, J. Martínez Sanz, M. Sánchez Conde, MJ. Vivancos, AM. Moreno, S.Serrano Villar, JL.Casado.

Hospital General Universitario Gregorio Marañón: PIs: J Berenguer, JC. López. Co-investigators: T. Aldamiz.

Hospital 12 de Octubre: PIs: D. Rial, R. Rubio. Co-investigators: F. Pulido, O. Bisbal, M. de Lagarde, A. Pinto, R. Font, O. Arce.

Hospital Universitario de La Princesa: PIs: I. de los Santos, J. Sanz. Co-investigators: LJ. García Fraile, A. Espiño, A. Sancha, M. Ciudad, A. Bautista, A. Gutiérrez.

Hospital Infanta Leonor-Vallecas: PIs: P. Ryan, G.Cuevas. Co-investigators: J.Troya, M.Matarranz, M.Torres Macho, J.Pardo-Guimera, J.Valencia, E.Jiménez.

Hospital Universitario Fundación Alcorcón: PIs:M. Velasco. Co-investigators: L. Moreno, R. Hervás, O. Martín, JE. Losa.

Hospital Universitario Príncipe de Asturias: PIs: J. Sanz Moreno. Co-investigators: M. Novella, C. Hernández, JA. Arranz, V Sampériz.

Hospital Universitario Rey Juan Carlos: PIs: S. Nistal.

Hospital General de Villalba: PIs: J. Hernández.

Hospital Universitario Infanta Elena: PIs: M. Rubio. Co-investigators A. Jiménez.

Hospital Quirón Salud: PIs: D.Carnevali. Co-investigators: J. Aguareles, A. Roda, P Guisado.

Hospital Universitario Infanta Sofía: PIs: I. Suárez.

Hospital Clínico San Carlos: PIs: J. Vergas, V. Estrada.

Hospital Puerta de Hierro: PIs: A. Díaz DE Santiago, S. de la Fuente.

Hospital Severo Ochoa-Leganés: PIs: M. Cervero

Hospital de Getafe: PIs: Sergio Rodriguez

Comunidad Foral de Navarra

Complejo Hospitalario de Navarra: PIs: M. Rivero, E.Moreno. Co-investigators: A. Arrondo, M Gracia, C. Ibero, J. Repáraz

Comunidad Valenciana

Hospital Clínico Universitario de Valencia: PIs: MJ. Galindo, R. Ferrando. Co-investigators: AI. De Gracia, M. García, N. Gómez, M. Martínez.

Hospital La Fé: PIs: M. Tasias, M. Montero. Co-investigators: M. Salavert, S. Cuéllar, E. Calbuig, M. Blanes, J. Fernández.

Hospital General Universitario de Valencia: PIs: M García del Toro.

Hospital General Universitario de Elche: PIs: F. Gutiérrez. Co-investigators: M. Masiá, S. Padilla, J. García, C. Robledano.

Hospital Marina Baixa: PIs: C. Amador

Hospital Universitario de la Plana: PIs: A. Blasco. Co-investigators: I. Pedrola, L. Fandos, M. Arnal.

Hospital General Universitario de Alicante:  PI: X. Portilla.

Hospital Arnau de Vilanova-Lliria: PIs: J. Flores, C. Tortaiada.

Hospital Peset: M. Madrazo, A. Artero.

Galicia

Hospital Álvaro Cunqueiro: PIs: G. Pousada, A. Ocampo. Co-investigators: M. Crespo, A. Pérez, C.Miralles.

Complejo Hospitalario Universitario de Ferrol: PIs: A. Mariño. Co-investigators: N. Valcarce, H.Alvarez, L. Vilariño, JF. García.

Hospital de Santiago de Compostela: A. Antela, E. Losada.

Islas Baleares

Hospital Universitari Son Espases: M. Riera, F. Alberti. Co-investigators: AM. Santos.

La Rioja

Hospital San Pedro – CIBIR: PIs: JR. Blanco. Co-investigators: V. Ibarra, J. Gallardo, JA. Oteo, L. Pérez.

País Vasco

Hospital de Donostia: PIs: JA. Iribarren MA, Von Wichmann. Co-investigators: X.Camino, MJ.Bustinduy, H.Azkune, M.Ibarguren, MA.Goenaga, I.Alvarez Rodriguez.

Hospital de Basurto: PIs: M.de la Peña, I Lombide. Co-investigators: M.López, J. de Miguel, V.Polo, S.Ibarra, OI.Ferrero, J.Muñóz.

Hospital Universitario Araba: PIs: JJ. Portu.

Hospital Osi Barakaldo-Sestao: PIs: R. Silvariño.
Hospital Universitario de Cruces: PIs: J. Goikoetxea, E.Bereciartua.

Principado de Asturias

Hospital Universitario Central de Asturias: PIs: V. Asensi, M. Rivas-Carmenado.

Murcia

Hospital Clínico Universitario Virgen de la Arrixaca: PIs: C. Galera. Co-investigators: M. Fernandez, H. Albendín, A.Castillo, MA.Merlos.

Hospital General Universitario Reina Sofía: PIs: E. Bernal. Co-investigators: T. Alcaraz.

**Appendix 2. Definition of comorbidities**

High blood pressure defined as systolic and diastolic values 130 and 85-89

Diabetes Mellitus defined as:

- Fasting blood glucose ≥126 mg / dl or,

- Blood glucose ≥200 mg / dl after oral glucose resistance test (2h) or,

- HbA1c ≥6.5%

Chronic kidney disease defined as:

- eGFR ≤ 60 mL / min for ≥ 3 months or,

- Albumin / creatinine ratio> 100 mg / mmol.

Chronic cardiovascular disease defined as the presence of any of the following in the clinical history: ischemic heart disease (myocardial infarction, angina pectoris), cardiopulmonary disease, arrhythmias and heart failure, cerebrovascular disease (hemorrhage, stroke, embolism, thrombosis, cerebral apoplexy or stroke), iseases of the arteries (atherosclerosis, aneurysm, embolism and arterial thrombosis).

Immune disorder (other than HIV infection) defined as the presence of any immunosuppressant/corticosteroid therapy in the clinical history
